# Response to Whole-Lung Low-Dose Radiation Therapy (LD-RT) Predicts Freedom from Intubation in Patients Receiving Dexamethasone and/or Remdesevir for COVID-19-Related Acute Respiratory Distress Syndrome (ARDS)

**DOI:** 10.1101/2021.02.10.21251242

**Authors:** Clayton B. Hess, Tony Y. Eng, Tahseen H. Nasti, Vishal R. Dhere, Troy J. Kleber, Jeffrey M. Switchenko, Brent D. Weinberg, Nadine Rouphael, Sibo Tian, Soumon Rudra, Luisa S. Taverna, Alvaro Perez, Rafi Ahmed, Mohammad K. Khan

## Abstract

**Background:** Phase I/II clinical trials have explored whole-lung low-dose radiotherapy (LD-RT) as a potential treatment for patients with COVID-19-related acute respiratory distress syndrome (ARDS). Initial findings require reproduction. Concomitant LD-RT administration with existing therapies requires safety evaluation.

**Methods:** Patients with COVID-19-related pneumonia receiving dexamethasone and/or remdesevir were treated with 1.5 Gy whole-lung LD-RT, followed for 28 days or until hospital discharge, and compared to controls blindly matched by age, comorbidity, and disease severity. Eligible patients were hospitalized, SARS-CoV-2 positive, had radiographic consolidations, and required supplemental oxygen. Endpoints included safety, clinical recovery, intubation, radiographic changes, and biomarker response.

**Findings:** 20 patients received whole-lung LD-RT between Jun 11 and Dec 7, 2020 and were compared to controls. Freedom from intubation improved from 68% in controls to 86% following LD-RT (p=0.09) as did C-reactive protein (CRP) (p=0.02) and creatine kinase (CK) (p<0.01) levels, consistent with prior report. Eighty percent of LD-RT patients experienced rapid decline in CRP within 3 days and were classified as LD-RT responders. Intubation-free survival (100% vs 66%, p=0.01) and oxygenation loads were lower in LD-RT responders compared to matched controls: 32% lower per individual (p=0.03) and 56% lower for the cohort (p=0.06). No patient whose CRP declined following LD-RT died or required intubation, whereas all LD-RT non-responders died. Observed reduction of prolonged recoveries and hospitalization times did not reach significance. Radiographic changes were equivalent.

**Interpretation:** A cohort of patients with COVID-19-related ARDS treated with LD-RT demonstrated superior freedom from intubation compared to matched controls, especially LD-RT responders (p=0.01). LD-RT appears safe to deliver with concurrent drugs. LD-RT lowered CRP and CK biomarkers. CRP response predicted favorable outcome. Optimal timing for LD-RT after oxygen dependence but before intubation may extinguish immunopathology prior to systemic spread. Confirmatory clinical trials are warranted. Clinical Trial Registration: NCT04366791.

**Funding:** None

## Introduction

The severe acute respiratory syndrome coronavirus 2 (SARS-CoV-2) and its viral syndrome, the coronavirus disease 2019 (COVID-19), have brought unprecedented death and global disruption. Mediated by a cascading, hyperinflammatory, macrophage-activating event in the lungs^1^, patients can face mortality rates of 30-50% once dependent on mechanical ventillation.^2-4^ The anti-inflammatory effects of low doses of ionizing radiation therapy (LD-RT) are well-known and have been shown to reduce inflammation in murine models of acute respiratory distress syndrome (ARDS) in the COVID-19 era.^5^ It has been hypothesized that LD-RT directed at the lungs may dampen cytokine hyperactivity through immunomodulation and improve outcomes of hospitalized patients.^6-8^ Initial reports of LD-RT have shown safety and potential benefit, meriting further investigation of this potential treatment for COVID-19.^9-11^ Yet, contradictory calls for caution and action demonstrate the barriers that remain and that further study is needed.^12-14^ Twenty-eight-day outcomes of two initial patient cohorts (n=10 combined) of a phase I/II American trial investigating safety and early efficacy of LD-RT for COVID-19 were previously reported.^9^ Outcomes of a confirmatory cohort of 20 additional COVID-19 patients treated with LD-RT are herein presented, compared with outcomes of 20 new controls blindly matched by age, comorbidity, and disease severity. The present cohort differs from prior in that all patients who received LD-RT also received concurrent dexamethasone and/or remdesevir, which has not been previously reported. This represents the first known confirmatory testing of initial trial observations following LD-RT for COVID-19-related ARDS and the first safety assessment of combined LD-RT with established COVID-directed drug therapies.

## Methods

### Trial Design

The Radiation Eliminates Storming Cytokines and Unchecked Edema as a 1-day Treatment for COVID-19 (RESCUE 1-19) trial is a 5-cohort, investigator-initiated, single-institution combined phase I/II trial. Cohort 1, comprised of 5 patients, was aimed to determine safety.^10^ Cohort 2, comprised of 5 additional patients, was designed to explore initial efficacy – combined outcomes from Cohorts 1 and 2 compared to 10 matched controls was previously reported.^9^ Cohort 3 explores LD-RT for intensive care unit patients and Cohort 5 explores a therapeutic dose reduction from 150 to 50 cGy – both are ongoing. The current manuscript reports outcomes from the completed Cohort 4, who received 1.5 Gy whole lung LD-RT with concurrent dexamethasone and/or remdesevir in oxygen-dependent patients with COVID-19-related ARDS. All participants gave written informed consent prior to any study procedures. Informed consent regarding potential late toxicities –risk of second cancer and accelerated cardiovascular disease -- was individualized to age and COVID-19-related mortality risk. Clinical Trial Registration Number was NCT04366791. The research protocol was approved by the Emory University Institutional Review Board.

A total of 20 hospitalized and oxygen-dependent patients received LD-RT and were followed for a minimum of 28 days or until discharge. Thereafter, a cohort of age- and comorbidity-matched controls was blindly and retroactively selected for comparative outcome analysis, using methods described previously.^9^ Twenty new controls were selected among SARS-CoV-2-positive patients who had previously enrolled on a separate, non-therapeutic, prospective institutional trial and matched by age, comorbidity, and disease severity. Study investigators were blinded to the selection and outcomes of control patients. Controls were permitted but not required to be co-enrolled on any therapeutic trial of COVID-19-directed drugs, including the Adaptive COVID-19 Treatment Trial (ACTT-1, Clinical Trial NCT04280705).

### Patients

Eligible LD-RT patients tested positive for SARS-CoV-2 by nasopharyngeal swab using polymerase chase reaction (PCR)-based testing, were hospitalized, had pneumonic consolidation on either chest radiograph (CXR) or computed tomographic (CT) imaging, and required oxygen supplementation. At the time of enrollment, exclusion criteria included disease severity that was too mild (on room air) or too severe (intubated). Patients who required oxygen levels beyond 15 L/min were considered transport ineligible. Anti-pyretic medications were suspended at enrollment. Following LD-RT, oxygen weaning followed hospital policy and standard of care. Patients underwent clinical assessment at the time of enrollment and on post-RT days 1, 3, 7, and 28, as well as optional assessment on days 14 and 21. Charlson Comorbidity Index (CCI)^15^ was used to assess comorbidity burden. Radiographs were permitted at any time as clinically indicated, but obtained per-protocol at least 12 hours prior to radiation, 24 hours following radiation, and on post-RT days 3, 7, 28, and optionally at days 14 and 21. Evaluation of serum inflammatory, renal, cardiac, chemistry, clotting, and hematologic markers were encouraged daily, but obtained at least at baseline and also on post-RT days 3, 7, 28, and optionally on days 14 and 21.

### Intervention

All enrolled patients received best supportive care plus dexamethasone and/or remdesevir prior to LD-RT delivery. They were subsequently administered a single treatment of 1.5 Gy to the bilateral whole lungs with 15 megavoltage photons on a linear accelerator, utilizing a 2-dimensional therapeutic radiation technique, an anterior-posterior beam configuration, and standard dose rates (600 MU/min). Patients in the control cohort received best supportive care and standard of care drug therapies for COVID-19 (ie, glucocorticosteroids, remdesevir, hydroxychloroquine, etc) per physician discretion or per separate therapeutic protocol, if enrolled. Convalescent plasma was not available for use. To match disease severity, eligible controls were required to have received supplemental oxygen during hospitalization and were excluded if they experienced rapid clinical decline and underwent intubation on the day of admission.

### Outcome Measures

Primary objectives were (1) to report the safety of LD-RT delivered concurrently with dexamethasone and/or remdesevir and (2) to investigate efficacy and reproducibility of signals observed in prior cohorts of the same clinical trial who received LD-RT without concurrent drug therapies.^9^ Efficacy was explored by comparing time to clinical recovery (TTCR). TTCR was defined equivalently as it was in the ACTT-1 trial and in prior cohorts, as the time from first COVID-19 intervention to the first day during which a subject satisfied one of three categories from an ordinal scale: (1) Not hospitalized, no limitations on activities; (2) Not hospitalized, limitation on activities and/or requiring home oxygen; or (3) Hospitalized, not requiring supplemental oxygen. A full 24-hour calendar day free of oxygen supplementation was required to trigger the binary classification of a patient as recovered. Additional secondary outcomes explored clinical course, radiographic changes, and clinical lab results. Clinical course was secondarily evaluated by overall survival (OS), total hospital duration, time from admission to clinical recovery, freedom from intubation, intubation-free survival, duration of intubation, and aggregated oxygen supplementation requirement. Intubation-free survival was defined as the proportion of patients who were both alive and had not required intubation or mechanical ventilation during hospitalization. Disease severity was assessed at baseline by oxygen requirement (L/min) and arterial blood gas using a ratio of arterial pressure (mmHg) of oxygen (PaO2) to fraction of inspired oxygen (FiO2) [P:F ratio]. Radiographic changes were evaluated by serial imaging. Chest radiographs were categorized as improved (I), stable (S), or worse (W) by comparing to the immediately preceding study by a blinded board-certified diagnostic radiologist (BW) and also blindly assigned an ordinal 1-5 score, using an acute respiratory distress syndrome (ARDS) scoring scale.^16^ Radiological blinding allowed knowledge of radiograph sequencing but ensured no knowledge of cohort designation, intervention received, or timing thereof. Chest computed tomography studies obtained at baseline and day 7 were subjectively assessed and visually compared without a standardized scoring system. Serological course was measured by serial laboratory evaluations of hematologic, renal, cardiac, chemistry, clotting, and inflammatory markers. Start time to clinical recovery (TTCR) was defined as it was in prior cohorts,^9^ as the time from LD-RT delivery (in the radiation cohort), as the first day of administration of COVID-19 therapy (in control patients, if received [n=18]), or as the first full-day of hospitalization (in control patients who received best supportive care alone [n=2]). To control for possible lead-time bias time resulting from this definition, time from hospital admission to clinical recovery was also evaluated. *LD-RT Response:* Following trial completion, staff reported observing consistent durable declines in C-reactive protein (CRP) following LD-RT in some patients, but persistent CRP rise in others that clinically seemed to correlate with outcome. Therefore, a predictive variable, *LD-RT response*, was retroactively created to investigate a predictive role of CRP response following LD-RT. Patients who did and did not experience consistent decline in CRP following LD-RT were categorized as *responders* and *non-responders*, respectively. LD-RT *non-responders* were defined by the following criteria: (1) elevated CRP level (>10mg/dL) the morning prior to LD-RT (baseline), and (2) two sequential rises in CRP above baseline among the first three daily measurements immediately following LD-RT. To control for lab variability, rise in CRP level was defined as an increase of ≥ 2% above the immediately preceding result. Daily labs were drawn approximately 24 hours apart so that duplicate or rapidly repeated lab rises did not overestimate sequential CRP rise. Evaluation was permitted to span 4 days following LD-RT in the event of a missing lab.

### Statistical Analysis

Wilcoxon rank sum tests and Fisher’s exact tests were used for continuous and categorical endpoints, respectively. For time-to-event endpoints, log-rank tests were used. Cumulative incidence of recovery and discharge were plotted for patients using the Kaplan-Meier method. Deceased patients were censored at time of death. Overall survival, freedom from intubation, and intubation-free survival were estimated using the Kaplan-Meier method. Univariate Cox proportional hazards models were fit and hazard ratios reported. Serial imaging ARDS scores were carried forward from Day 7 to 14 to 21 if missing. CXR outcomes were reported as mean ARDS scale scores for sequential time periods.

Median and interquartile range was calculated for laboratory values at clustered time points: hospital days 0-2, 3-5, 6-8, 9-11, and 12-14. Univariate ANOVA (within clustered days) and multivariate repeated values ANOVA (between clustered days) were used to compare changes in repeated laboratory measures between cohorts on, respectively. Clinical outcomes were stratified by binary classification of patients as *responders* vs. *non-responders* based on CRP response following LD-RT in an unplanned analysis for hypothesis generation based on observation made during the trial. Within this subset analysis, t-tests were used for continuous endpoints rather than Wilcoxon rank sum tests because of the observation of effect specifically on outlying data points likely to affect data means rather than medians. Means were reported wherever medians did not explain cohort differences. Statistical analysis was performed using SAS 9.4 (SAS Institute Inc., Cary, NC), and statistical significance was two-sided and assessed at the 0.05 level.

## Results

### Patients

From June 11, 2020 to December 7, 2020, 65 patients were screened for eligibility, and twenty-five were enrolled. Of these, the first twenty were treated with LD-RT. The last five were enrolled as serologic controls for study of immune marker correlates and did not receive LD-RT (serological data to be reported at a later date). None of the 20 enrolled patients required more than 15 L/min of supplemental oxygen, making all patients eligible for transport to the linear accelerator for LD-RT delivery. Of the 40 patients who were screened but not enrolled, 21 had disease too mild to meet severity criteria, 5 declined to participate, 5 were young and likely to spontaneously recover, 5 had no documented rationale for non-enrollment, 3 had disease too severe to meet severity criteria and were intubated before written consent, and 1 did not ultimately meet SARS-CoV-2 eligibility criteria (Figure 1). Following trial completion, 20 separate patients who had been hospitalized between March 6, 2020 and November 19, 2020 were blindly matched. One was found to have rapidly intubated on the day of admission, was excluded, and then blindly replaced to ensure cohort matching of disease severity.

**Figure 1.**
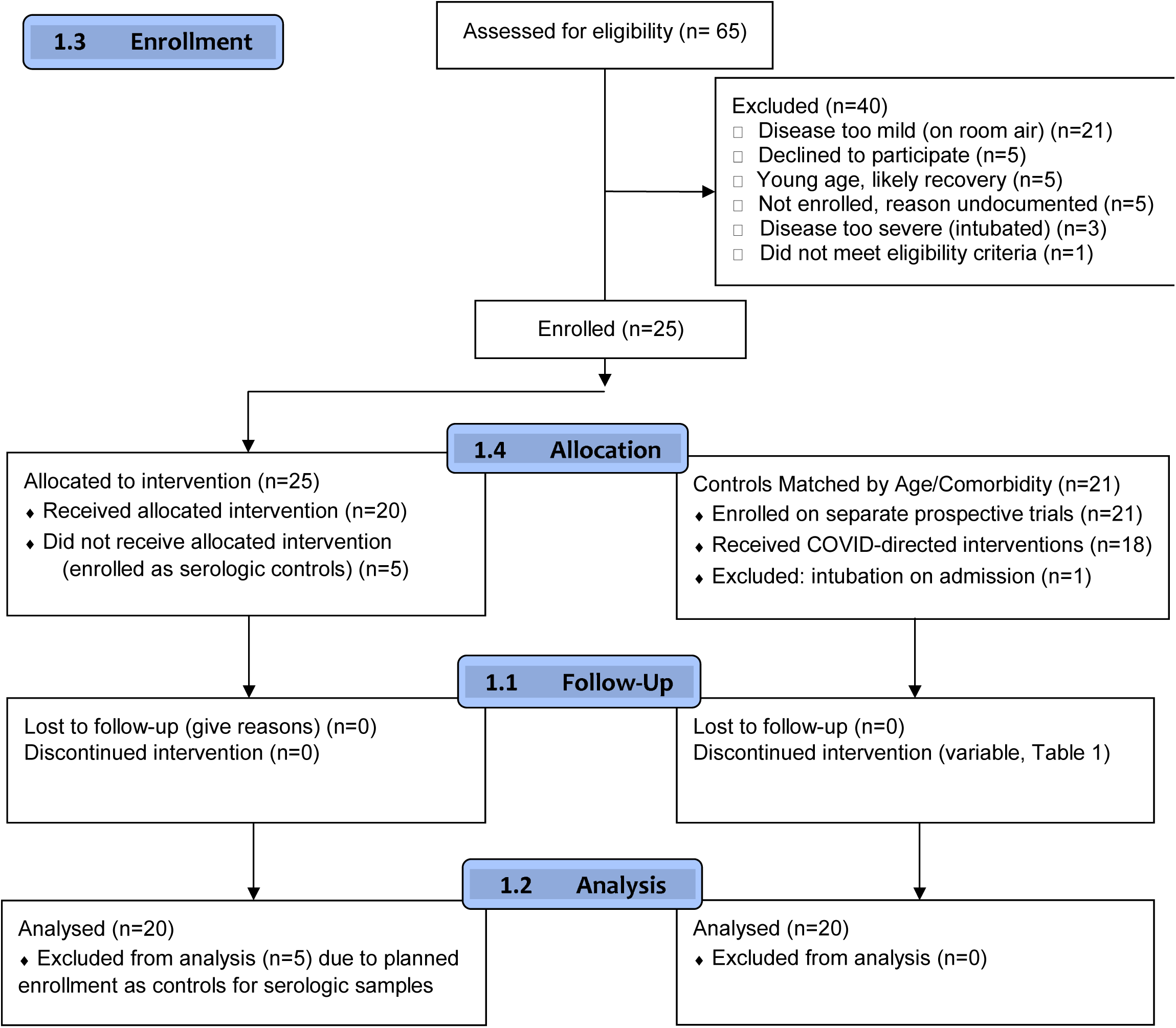
CONSORT Flow Diagram.

Table 1 outlines well-balanced patient demographics at the time of hospital admission and administered COVID-19 drug therapies. Median age was 63 (49-88). Fifty-three percent of patients were African-American, 38% were female, and 12% were residents of assisted living centers. No patients experienced altered mentation at enrollment. Median CCI comorbidity scores were: 3 (range 1-12). Median duration of symptoms prior to admission was 10 (range 1-15) and 7 (2-14) days following LD-RT and for controls, respectively (p=0.10). Median oxygen supplementation requirement at the time of admission was 3 liters (range 0-15) for both cohorts (p=0.51). Disease severity, as assessed by median ratio of arterial pressure (mmHg) of oxygen (PaO2) to fraction of inspired oxygen (FiO2) [P:F ratio] was not statistically different between the cohorts: 169 (range 122-325) compared to 183 (range 94-314), respectively (p=0.84). Patients in both the LD-RT and control cohorts received COVID-directed drug therapy for a median of 7.5 days (p=0.87). LD-RT was administered at median hospital day 3 (range 1-8). Patients in the LD-RT cohort received COVID-directed therapy (dexamethasone and/or remdesevir, etc.) at a median start time of hospital day 1 (range 1-5), compared to 2.5 (range 1-6) in controls (p=0.02).

**Table 1.**
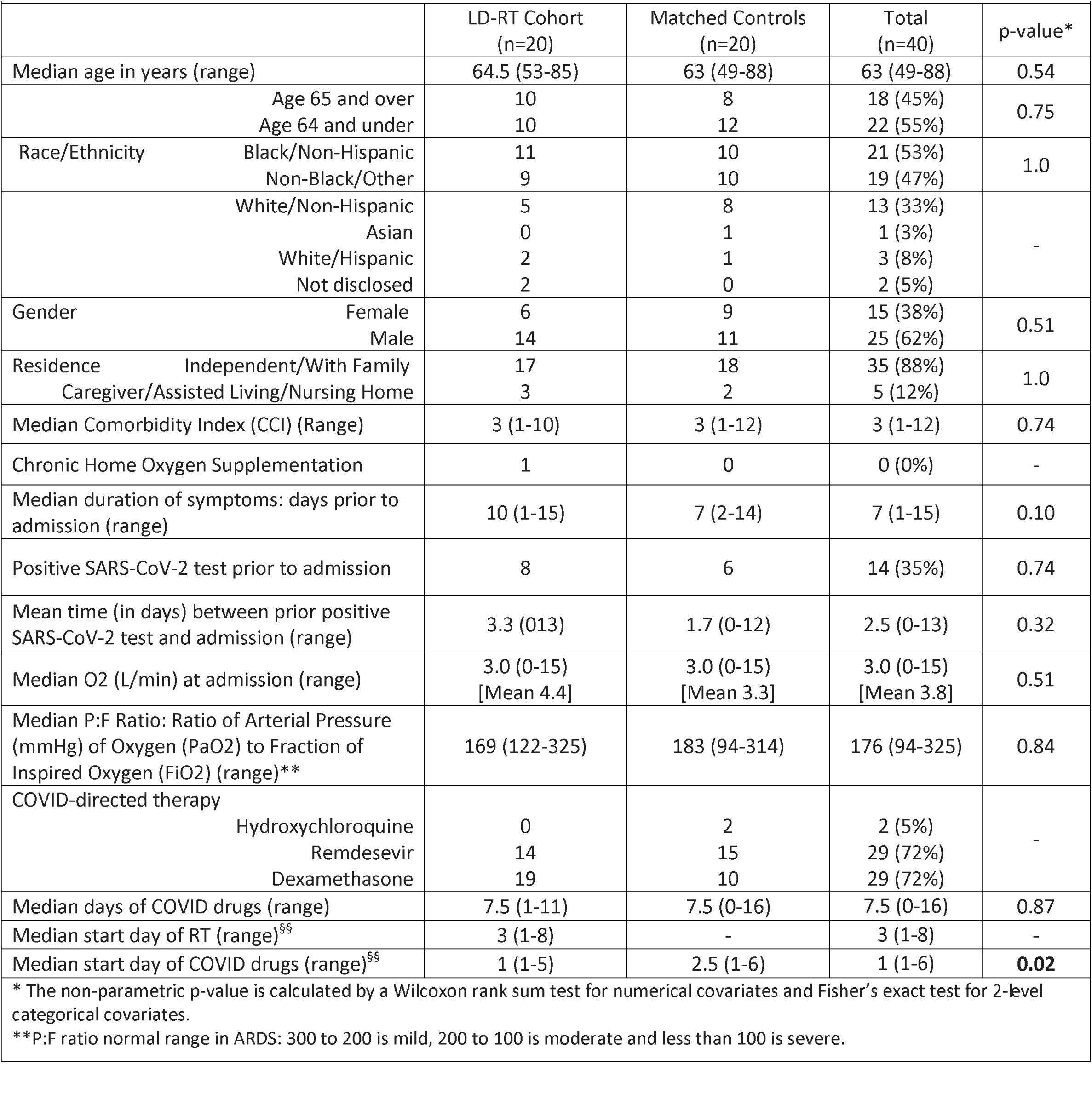
Patient demographics

### Clinical Outcomes

LD-RT was delivered safely and without acute toxicity or reflex clinical, radiographic, or serologic worsening of ARDS when administered concurrently with dexamethasone and/or remdesevir. Median time to clinical recovery (TTCR) and time from admission to clinical recovery were not statistically different between the cohorts (p=0.37 for both, Table 2). Freedom from intubation at day 28 was 86% following LD-RT compared to 68% in controls (p=0.09) (Figure 2), and intubation-free survival was 77% compared to 68%, respectively (p=0.17). Median hospital duration was 10.5 days (range 5-33) in the LD-RT cohort compared to 11.5 days (range 3-42) in controls (p=0.61). Additional treatment outcomes for the entire cohort are reported in Table 2.

**Table 2.** Day 28 Outcomes for the Entire LD-RT Cohort vs. Matched Controls (n=40)

|  |  |  |  |  |  |  |
| --- | --- | --- | --- | --- | --- | --- |
| Variables |  | LD-RT Cohort (N=20) |  | Control Cohort (N=20) |  | p-value* |
| Categorical | Outcome | Yes | No | Yes | No |  |
|  | Clinically Recovered | 16 | 4 | 17 | 3 | 1.0 |
|  | Mechanically Ventilated | 2 | 18 | 6 | 14 | 0.24 |
|  | Deceased** | 3 | 17 | 0 | 20 | 0.23 |
| Continuous |  | Median | Range | Median | Range |  |
|  | Hospital Duration (days) | 10.5<br>Mean 12.5 | 5-33 | 11.5<br>Mean 15.8 | 3-42 | 0.61 |
|  | Duration of Oxygen Supplementation (days) | 10.5<br>Mean 12.1 | 4-33 | 9.5<br>Mean 13.9 | 3-42 | 0.72 |
|  | Duration of Ventilation, if intubated (days) | 9.5<br>Mean 9.5 | 5-14 | 9.5<br>Mean 13 | 1-34 | 0.87 |
|  | Average Daily Maximum Oxygen Need (L/min) | 6.7<br>Mean 15.3 | 1.5-81.5 | 4.0<br>Mean 23.1 | 1.5-86 | 0.91 |
|  | Cumulative Cohort Oxygenation Need<br>(max daily L/min * days oxygenated) | 48.3<br>Mean 210 | 6-1591 | 60.3<br>Mean 530 | 7.5-3636 | 0.73 |
|  | Days Febrile | 1.0<br>Mean 2.0 | 0-9 | 1.0<br>Mean 3.0 | 0-12 | 0.91 |
| Time-to-Event |  | Median | Range | Median | Range |  |
|  | Time to Clinical Recovery [TTCR] (days) | 7.5 | 2-30 | 7 | 2-38 | 0.37 |
|  | Time from Admission to Clinical Recovery (days) | 10 | 4-33 | 9 | 2-41 | 0.37 |
|  | Freedom from Intubation at day 28 (percent, 95% CI) | Not reached | 86 (54-97) | Not reached | 68 (42-84) | 0.09 |
|  | Intubation-free Survival at day 28 (percent, 95% CI) | Not reached | 77 (42-92) | Not reached | 68 (42-84) | 0.17 |

\* The non-parametric p-value is calculated by a Wilcoxon rank sum test for numerical covariates, Fisher's exact test for 2-level categorical covariates, and log-rank test for time-to-event covariates with medians reported from Kaplan-Meier curves.
\*\* (1) Patient around 80 to 85 years old with do-not-resuscitate orders, whose family refused intubation, de-escalated care, and pursued comfort measures. Died hospital day 11, hypoxic after refusing high-flow oxygen support. (2) Patient around 55 to 60 years old who received dexamethasone and LD-RT before intubation. Died on hospital day 8 despite aggressive care. (3) Patient around age 70 to 75 who received dexamethasone and LD-RT before intubation. Died on hospital day 25 despite aggressive care.

**Figure 2.**
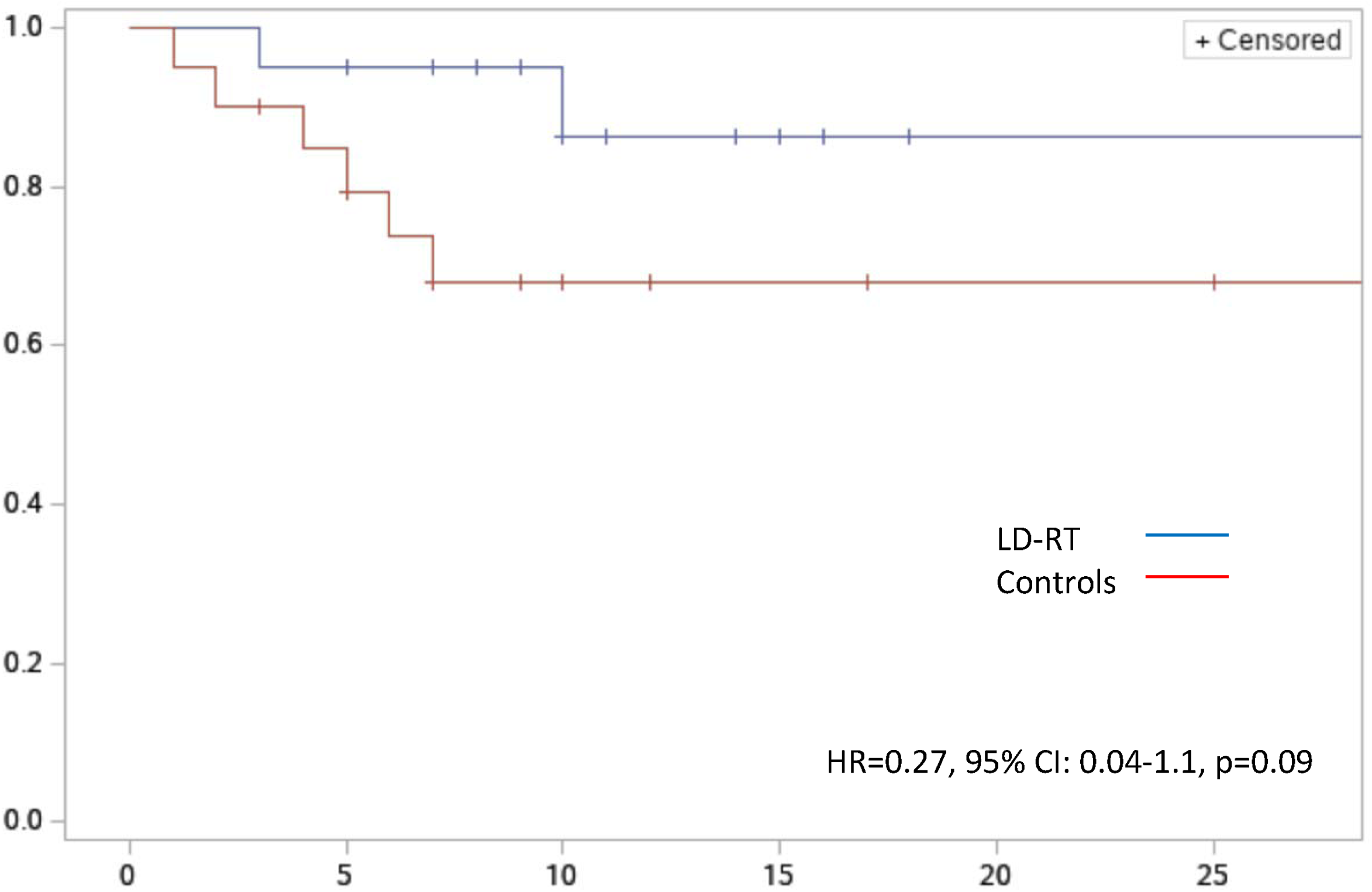
Freedom from Intubation for the Entire LD-RT Cohort vs. Matched Controls (n=40)

### Radiographic Response

Radiographic improvement was equal between the LD-RT and control cohorts (p=0.72, Figure S1). Representative CT images from patients with high burden of pulmonary consolidations associated with COVID-19 that radiographically worsened following LD-RT, as well as an example of radiographic improvement, are shown in Figure S2.

### Serologic Response

Hematologic, renal, cardiac, chemistry, clotting, and inflammatory markers were tracked from the first day of hospitalization. Plotted medians and inter-quartile ranges of serologic biomarkers for both LD-RT and control patients are shown in Figure S3. Decline of inflammatory biomarker C-reactive protein (CRP) was statistically superior in the LD-RT cohort compared with controls (p=0.02). Cardiac marker, creatine kinase, started higher in LD-RT patients, but was also significantly reduced over controls (p<0.01). Lower troponin-1 levels in the LD-RT were not statistically detectable (p=0.29). Liver function remained normal following LD-RT, while transaminitis occurred in some controls, but did not reach significance (AST p=0.35; ALT p=0.60). Neutrophil-to-lymphocyte ratio trended higher following LD-RT (p=0.12). There were no detectable reductions (p=0.80) in white blood cell count. No neutropenia was observed. Renal biomarker creatinine was not significantly affected (p=0.75). Myoglobin, erythrocyte sedimentation rate, lactate dehydrogenase, monocyte count, ferritin, fibrinogen, procalcitonin, and interleukein-6 trended downward after LD-RT but did not reach significance or control comparisons were not different or not available (Figure S3).

### LD-RT Responders

Sixteen of 20 irradiated patients (80%) experienced rapid decline in CRP levels over ≤3 days following LD-RT and were classified as *LD-RT responders*. The remaining four (20%) experienced rise in CRP and were classified as *non-responders (Figure S3, bottom right pane)*. Outcomes for these 16 LD-RT responders compared with their 16 matched controls are shown in Figure 3 and Table 3 (n=32). Time to clinical recovery was 7 days (range 2-15) in LD-RT responders and 7 days (range 3-38) in controls with observed reduction in prolonged recoveries seen as upper curve separation (p=0.29, Figure 3a). Median time from admission to clinical recovery was 9.5 days (range 1-17) following LD-RT compared to 9 days (range 1-41) in controls, with the same upper curve separation (p=0.26, Figure 3b). Median hospital duration was 10 days (range 5-18) for LD-RT responders compared to 10 days (range 3-42) for controls, which also showed fewer prolonged hospitalization and upper curve separation (p=0.22, Figure 3c). As a categorical variable, none of the LD-RT responders died or required intubation, compared to 5 of 16 matched controls (p=0.04, Table 3). As a time-to-event endpoint, median intubation-free survival was not reached in either cohort, but overall freedom from intubation were 100% for LD-RT responders and 66% for matched controls (p=0.01, Table 3, Figure 3d). Median duration of time intubated was 0 days for LD-RT responders vs. 10 days in matched controls who required intubation. Mean total time requiring oxygen supplementation prior to recovery was 10.5 days (range 4-18) in LD-RT responders compared to 14.3 days (range 3-42) in controls (p=0.24). Average daily oxygen maximums per patient were 32% lower at 7.6 L/min (mean) in LD-RT responders compared to 24 L/min in controls (p=0.03, Table 3). For the cohorts cumulatively, mean aggregated amount of oxygen supplementation (maximum daily L/min x total days oxygenated per patient) was 56% lower at 94.2 vs. 593 L/min*days, respectively (p=0.06). Average number of days febrile (any fever) was 1.4 days (range 0-9) vs. 3.3 days (range 0-12), respectively (p=0.14, Table 3), in favor of LD-RT responders. Additional treatment outcomes for LD-RT responders are reported in Table 3. Of 4 non-responders, 2 required intubation, 3 died by day 28 (1 refused intubation), and the 4th died on day 33.

**Table 3.** Day 28 Outcomes for LD-RT Responders vs. Matched Controls (n=32)

| Variables |  | LD-RT Responders (N=16) |  | Control Cohort (N=16) |  | p-value* |
| --- | --- | --- | --- | --- | --- | --- |
| Categorical | Outcome | Yes | No | Yes | No |  |
|  | Clinically Recovered | 16 | 0 | 13 | 3 | 0.23 |
|  | Mechanically Ventilated | 0 | 16 | 5 | 11 | **0.04** |
|  | Deceased | 0 | 16 | 0 | 16 | 1.0 |
| Continuous |  | Median | Range | Median | Range |  |
|  | Hospital Duration (days) | 9.5<br>Mean 10.8 | 5-**18** | 9.5<br>Mean 15.5 | 3-42 | 0.15 |
|  | Duration of Oxygen Supplementation (days) | 9.5<br>Mean 10.5 | 4-**18** | 9.5<br>Mean 14.3 | 3-42 | 0.24 |
|  | Duration of Ventilation, if intubated (days) | 0 | NA | 10<br>Mean 14.2 | 1-34 | - |
|  | Average Daily Maximum Oxygen Need (L/min) | 4.8<br>Mean 7.6 | 1.5-**30** | 7.1<br>Mean 24.0 | 1.5-86 | **0.03**<br>**32% lower** |
|  | Cumulative Cohort Oxygenation Need<br>(max daily L/min * days oxygenated) | 46.2<br>Mean 94.2 | 6-**454** | 104.5<br>Mean 593 | 8-3636 | **0.06**<br>**56% lower** |
|  | Days Febrile | 1.0<br>Mean 1.4 | 0-9 | 1.0<br>Mean 3.3 | 0-12 | 0.14 |
| Time-to-Event |  | Median | Range | Median | Range |  |
|  | Time to Clinical Recovery (days) | 7 | 2-**15** | 7 | 3-38 | 0.29 |
|  | Time from Admission to Clinical Recovery (days) | 9.5 | 4-**17** | 9 | 2-41 | 0.26 |
|  | Freedom from Intubation at day 28 (percent, 95% CI) | Not reached | 100% | Not reached | 66 (36-84) | **0.01** |
|  | Intubation-free Survival at day 28 (percent, 95% CI) | Not reached | 100% | Not reached | 66 (36-84) | **0.01** |

\* The parametric p-value is calculated by a t-test for numerical covariates, Fisher's exact test for 2-level categorical covariates, and log-rank test for time-to-event covariates with medians reported from Kaplan-Meier curves. Notable range reductions and statistical trends in **bold**.

**Figure 3a.**
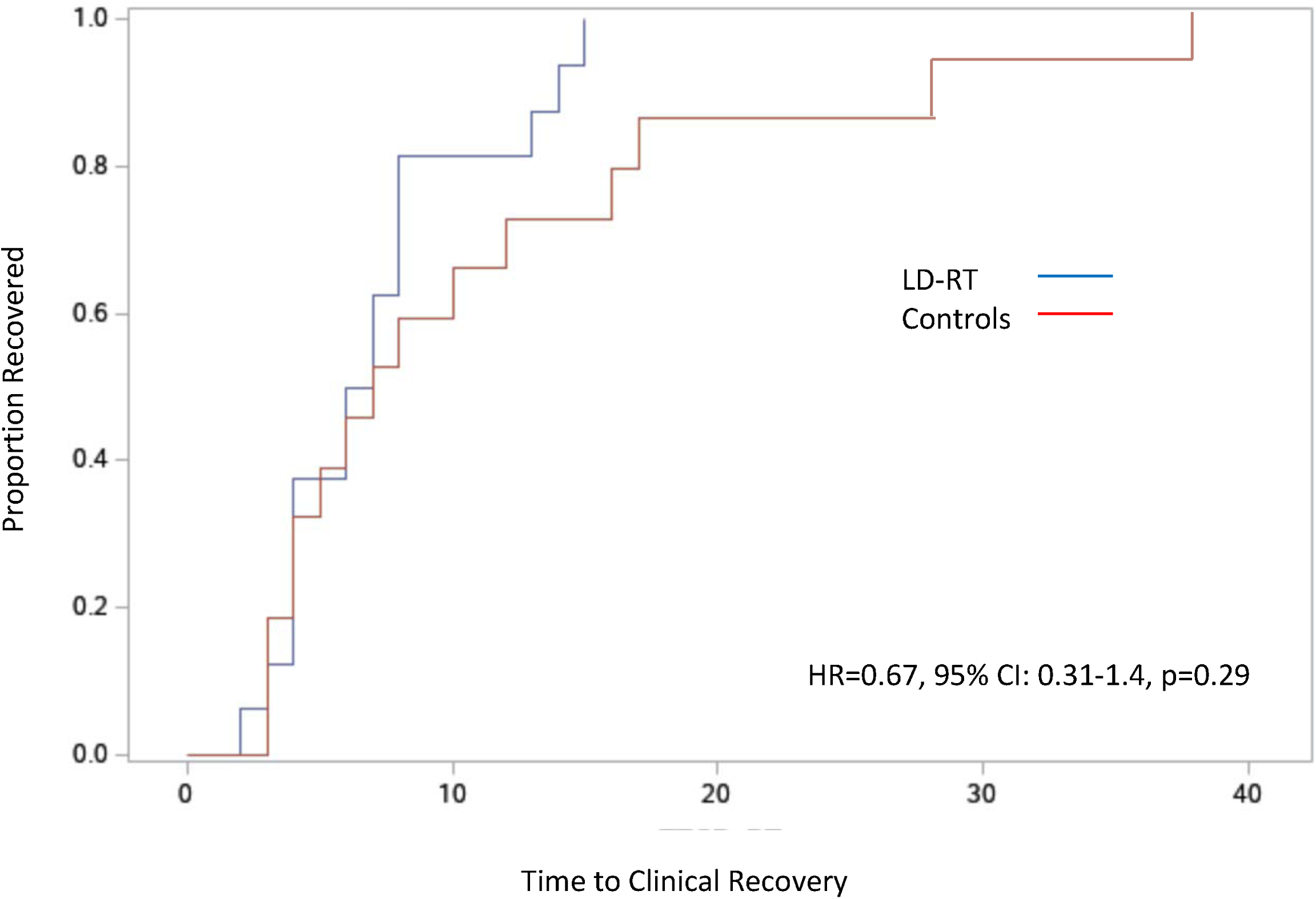
Time to Clinical Recovery for LD-RT responders vs. controls (n=32) *Defined from LD-RT (top) or first day of COVID drug in controls (bottom)

**Figure 3b.**
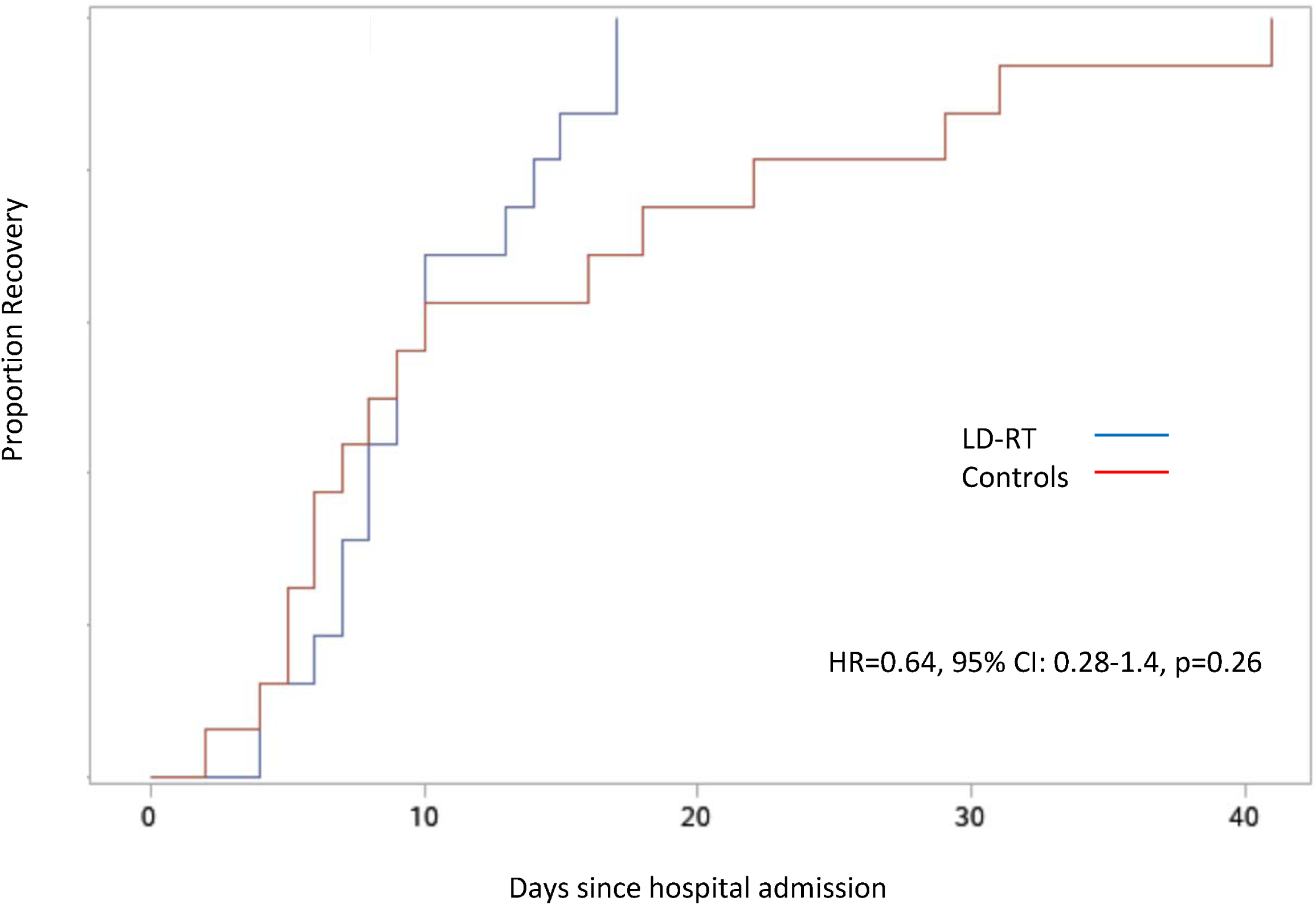
Time from admission to clinical recovery for LD-RT responders vs. controls (n=32)

**Figure 3c.**
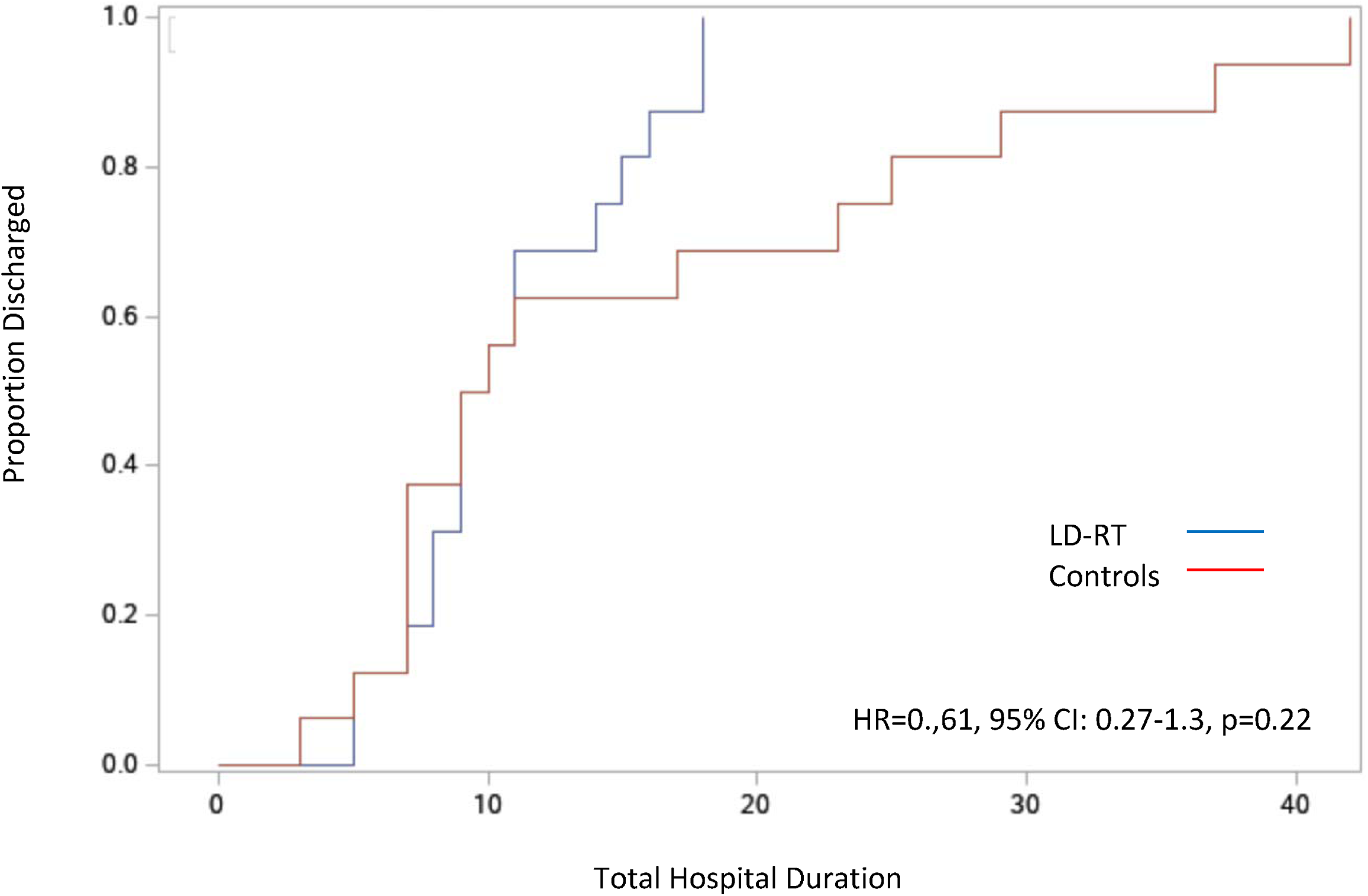
Total hospital duration for LD-RT responders vs. controls (n=32)

**Figure 3d.**
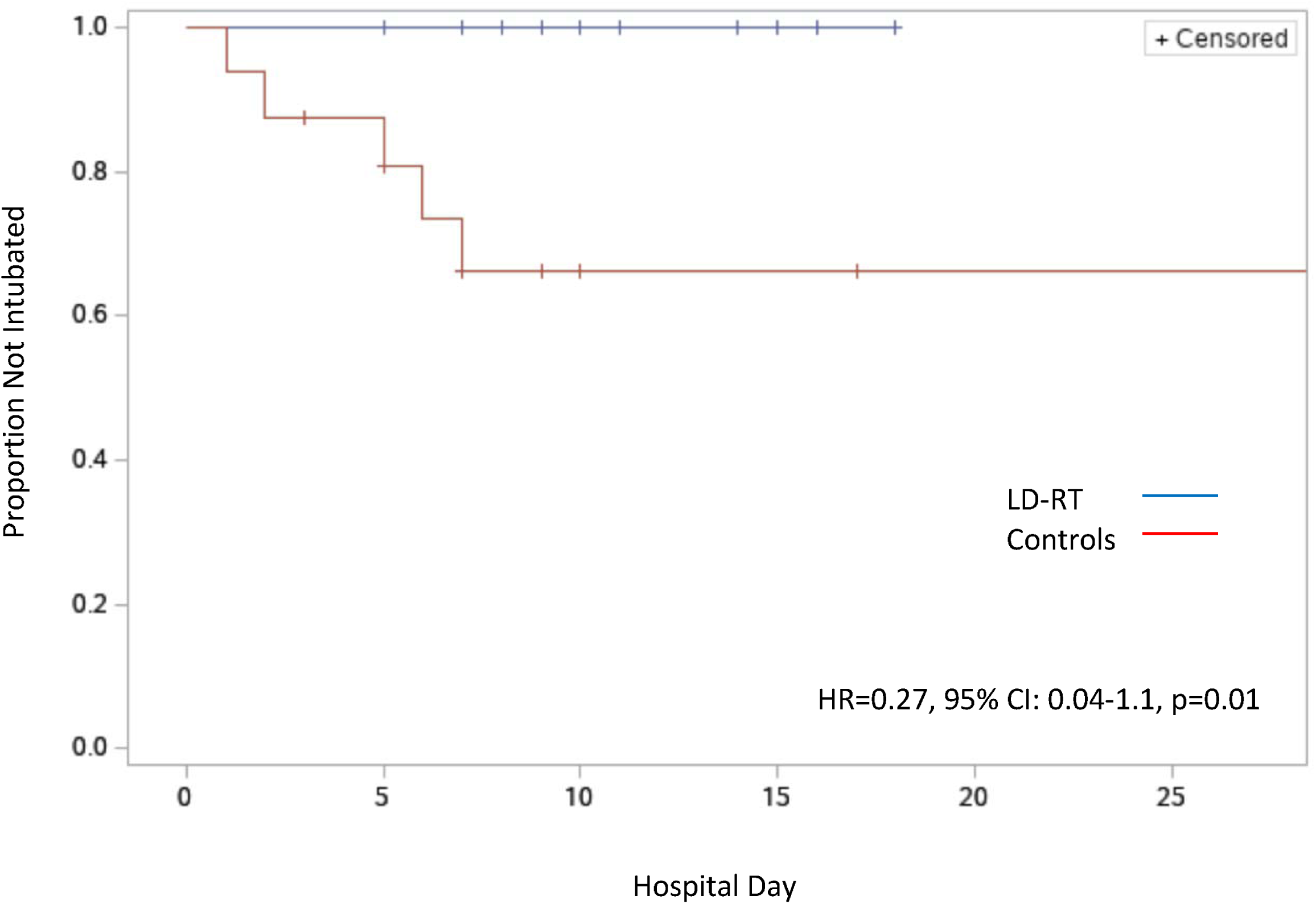
Intubation-free survival for LD-RT responders vs. controls (n=32)

### Adverse Events

There were no recorded acute toxicities. No LD-RT patients experienced rapid reflex worsening of symptoms, radiographs, or serologies nor was there compounding immunosuppression or cytopenias when LD-RT was given concurrently with dexamethasone. The 3 deaths observed at day 28 in the LD-RT cohort are described in Table 2. Two patients had COVID-19 symptoms escalate despite LD-RT; both went on to intubation and could not be weaned. The third was a patient with a large brain meningioma whose family removed all supportive care and refused intubation and who died hypoxic after removing high-flow oxygen. Contrary to expectations based on known mortality rates, none of the 20 blindly selected controls died. No other toxicity, airway emergencies, or adverse events were observed following LD-RT.

## Discussion

This report describes 28-day outcomes of the first phase II trial in humans exploring efficacy of immunomodulatory, whole-lung, low-dose radiation therapy (LD-RT) given concurrently with dexamethasone and/or remdesevir for patients with COVID-19-related ARDS. Cohorts were well matched without significant differences in confounding variables (Table 1). Concurrent delivery of LD-RT together with COVID-directed therapies appears safe.

A primary goal of this trial was to explore efficacy and to attempt to reproduce prior efficacy signals.^9^ Among these, the hypothesis that LD-RT prevents intubation was corroborated. Freedom from mechanical ventilation was improved following LD-RT compared to controls, from 68% to 86% in the entire cohort (p=0.09, Figure 2). Among LD-RT responders, whose CRP rapidly declined following LD-RT, intubation was not required at all, while it was still needed in 31% of matched controls. In prior cohorts, freedom from intubation improved from 60% to 90% following LD-RT.^9^ Given the reproducibility of this finding in 20 new patients, the hypothesis that LD-RT may lower intubation risk in COVID-19 is further strengthened, especially in LD-RT responders. If true, LD-RT’s potential reduction in total oxygenation days, days intubated, hospital duration, and the resources required to support patients with COVID-19-related ARDS may be quite dramatic. Due to this finding, the endpoint of an ongoing randomized trial has been revised to power the study for intubation-free survival and a reduction for the need of mechanical ventilation.

We also reproduced important serologic responses, wherein elevations in C-reactive protein (p=0.02) and creatine kinase (p<0.01) improved following LD-RT, even in the presence of dexamethasone. This supports the hypothesis that pulmonary hyper-inflammation can be extinguished upstream to avoid downstream effects on other organs, like the heart. A rapid decline in CRP following LD-RT may be indicative that hyper-inflammatory cascades may be localized within the lungs. Trends continue to suggest that LD-RT may have a protective effect in preventing elevations in other cardiac markers (troponin-1), hepatic injury, as well as coagulopathies, but these values did not reach significance and require additional study. Notable findings from Cohort 1 and 2 that could not be reproduced, include radiographic improvement compared to controls (Figure 3 and Figure S1). Radiographic recovery may occur over a longer timeline, as previously reported.^17^

While neither median time to clinical recovery (TTCR) nor the median time between admission and clinical recovery were statistically different between the cohorts overall, we performed a subset analysis of the 80% of patients with rapid serologic response to LD-RT, based on observations made during the trial. We noted that CRP response immediately following LD-RT seemed to predict clinical outcome and that few such patients went on to experience prolonged intubation. This subset analysis defined LD-RT responders as those patients whose CRP values rapidly declined following LD-RT. What we have observed supports effect modification, wherein outcomes following LD-RT differ by whether there is rapid serologic response in CRP levels. LD-RT responders followed clinical trends similar to what was observed in Cohorts 1 and 2, with fewer prolonged recoveries and shorter hospital durations (Figure 3a-c).^9^ In fact, no one who had a rapid decline in CRP went on to intubate or die. Non-responders did poorly, even worse than matched controls. This suggests that LD-RT may distinguish localized from systemic spread of the “cytokine storm.” This predicts that LD-RT responders (localized COVID) and non-responders (systemic COVID) would, respectively, have superior and inferior outcomes to matched controls not stratified by localization of immunopathology. In retrospect, the designation of LD-RT responder as a variable was also predictive of outcome in patients treated in Cohorts 1 and 2. The only patient who intubated/died following LD-RT in that cohort had persistent rise in CRP over 3 days (see reference supplemental materials, Cohort 1, patient 5).^9^ All other patients had rapid CRP response after LDRT and recovered without intubation.

Metaphors comparing COVID-19 to a brushfire have become useful among trial staff for patient education, wherein the ability for LD-RT to stop COVID-19’s cytokine storm in the lungs is compared to firehoses used to put out a small fire before it can spread into a raging wildfire. The time required to turn the CRP around within the lungs (anecdotally by day 3) has been compared to the distance required to stop the momentum of a speeding firetruck. Among the 30 patients who received LD-RT in cohorts, 1, 2 and 4, 25 (83%) had their CRP rapidly decline after treatment – of these, none went on to intubation or death. The other 5 experienced either no slowing of CRP rises at all or only a transient one-day dip before further rises (Figure S3, bottom right panel). In these, COVID-19’s fire-like “cytokine storm” may have already spread beyond the lungs and whole-lung LD-RT may have been too little delivered too late to prevent systemic spread of the infection. As CRP is among the most potent biomarker predictors of mortality and outcomes in COVID-19, the rapid and reproducible decline in CRP seen following LD-RT merits additional investigation.^18^

There are important differences between outcomes from the present cohort compared with cohorts 1 and 2 that may aid in understanding why the cohort collectively did not demonstrate improvement in time to clinical recovery as was seen previously. First, triggering of TTCR required a 24-hour period off of oxygen in cohort 4, compared to 12 hours in prior cohorts. This definition revision was due to mimicry of the ACTT-1 trial, which changed its definition at the time of publication, as discussed previously.^9^ Second, our population of enrolled patients changed as time progressed throughout 2020. Whereas our initial population was comprised of mostly elderly individuals, by July 2020, nursing homes in our area were no longer experiencing major outbreaks. Cohorts 1 & 2 had much older patients with milder disease, and the current study had younger patients requiring higher liters/min of oxygen supplementation. Our experience suggests that older patients may have had more rapid response to LD-RT compared to younger patients with higher burden of disease. Third, LD-RT was delivered later during hospitalization in prior cohorts and earlier (median day 3) in the present study. Fourth, dexamethasone and remdesevir were given concurrently in the present study but not in prior cohorts, which may have reduced the observable difference between cohorts. The neutrophil-to-lymphocyte ratio was lower following LD-RT in initial cohorts, but higher in the present study with a trend towards significance (p=0.12). This lab result has been correlated with inferior outcomes and the inversion of the finding between cohorts merits further investigation.^19^ This may be due to the improbable favorability of outcomes of blindly selected controls (0% mortality) and the high proportion (95%) of dexamethasone use (causes leukocytosis) in patients receiving LD-RT (causes lymphopenia), compared to controls (50%, Table 1).

While we have controlled for prior study weaknesses including variations in recovery definitions and transport eligibility, certain limitations remain. These include a non-randomized approach, exploratory intent, small patient numbers, differing control treatments, different laboratory and imaging schedules between the LD-RT and control cohorts, variable times of symptom onset, limited imaging (Figure S2) and serological (Figure S3) studies in the control cohort before intervention and beyond 7 days, and lack of detailed viral load evaluations in the LD-RT and control cohorts. Future work with LD-RT will include confirmatory testing for efficacy using a phase III design, exploration of lower dose ranges, detailed CD-8 T-cell activation studies, CD-4 T cell activation, changes in B-cell profiles, antibody formation, cytokine analysis, and neutralization tests.

In summary, this report corroborates prior finding that LD-RT may prevent intubation in COVID-19. Our results show that LD-RT again demonstrated an anti-inflammatory effect in COVID-19 that may lower the risk of mechanical ventilation, the duration of oxygen supplementation, and total hospital recovery time, especially in patients who experience rapid decline in CRP following LD-RT. LD-RT again reduced CRP and CK levels, denoting cardiac protection. Early LD-RT may be protective against downstream elevations in troponin-1, D-dimer, hepatic injury, and cardiac injury. Reduction of CRP following LD-RT may be indicative of localized inflammation. Further studies of LD-RT are needed in patients with COVID-19 to prove efficacy and further inform its optimal role in the exiting COVID-19 treatment landscape. As of January 2021, more than 95 million people globally are confirmed as infected with SARS-CoV-2, leading to over 2,032,000 known COVID-19-related deaths. The worldwide implications of LD-RT as a rapid, inexpensive, and globally-available treatment cannot be dismissed, especially given this second confirmatory cohort showing reproducibility of lower rates of intubation. Phase III randomized trials are ongoing, but the speed of these investigation could be accelerated with additional efforts by the radiation oncology community.

## Conclusion

A hospitalized cohort of patients with COVID-19 received LD-RT concurrently with dexamethasone with or without remdesevir and showed a trend towards intubation prevention (p=0.09). The addition of LD-RT to drug therapies appears safe. Subset analysis revealed that patients whose CRP rapidly declines following LD-RT did not die or require intubation. This may be indicative of localized ARDS disease process within the lungs and prognostic of favorable outcome. LD-RT responders did not require intubation and had lower burden of oxygen requirements with trending reductions in recovery and hospitalization times. Biomarkers CRP and CK responded to LD-RT, corroborating anti-inflammatory and cardio-protective effects. Prevention of downstream coagulopathy and hepatic dysfunction remain feasible. Early intervention after oxygen dependency but prior to intubation may be an optimal therapeutic timeline. Further clinical trials are justified [Clinical Trial Registration: NCT04433949]

## Data Availability

Data can be provided upon appropriate request and approval by the PI of the trial and per institutional approval processes.

## Figure Legends

**Figure S1.**
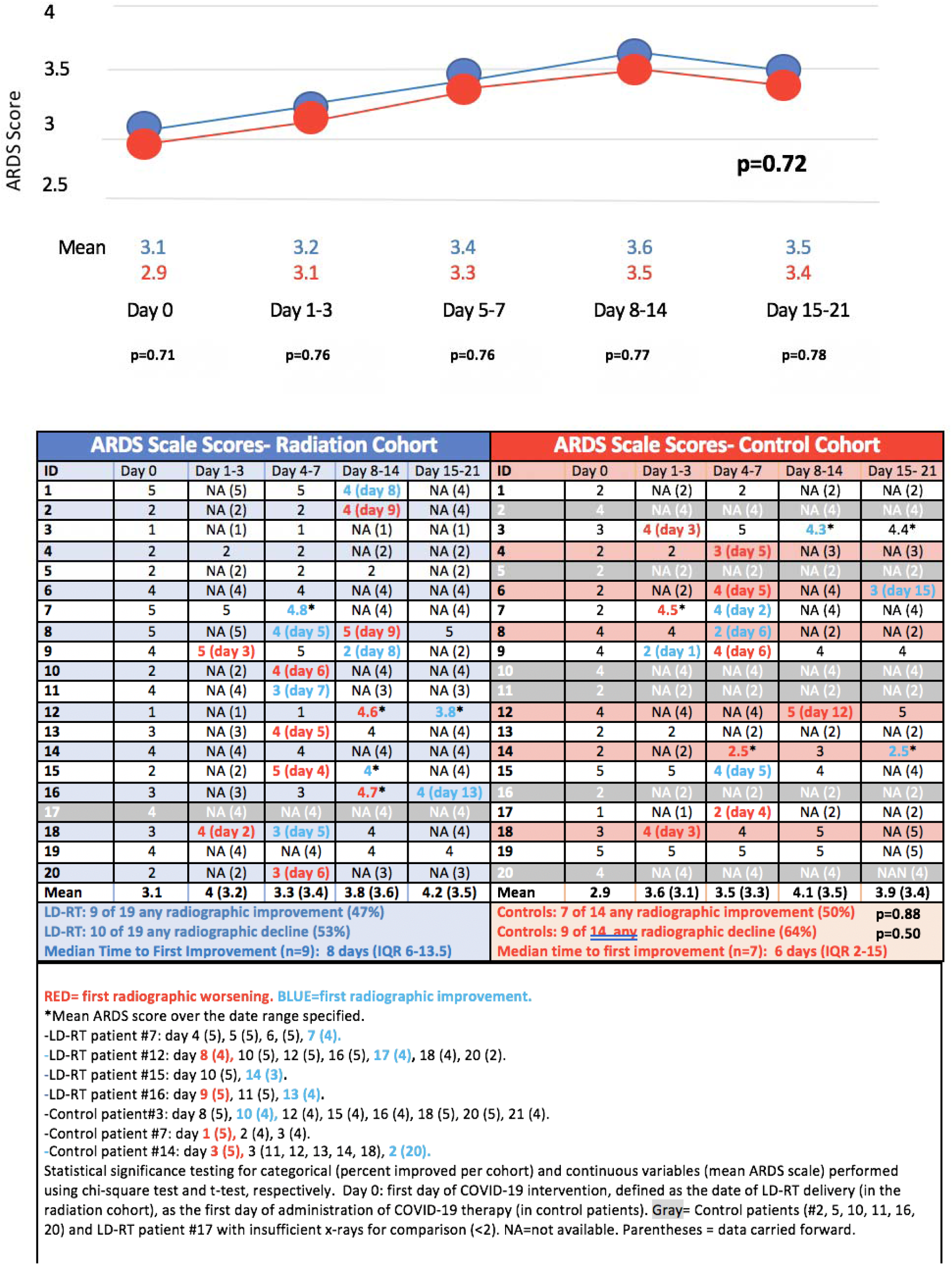
ARDS X-ray scale scores Pre- and post-COVID directed intervention.

**Figure S2.**
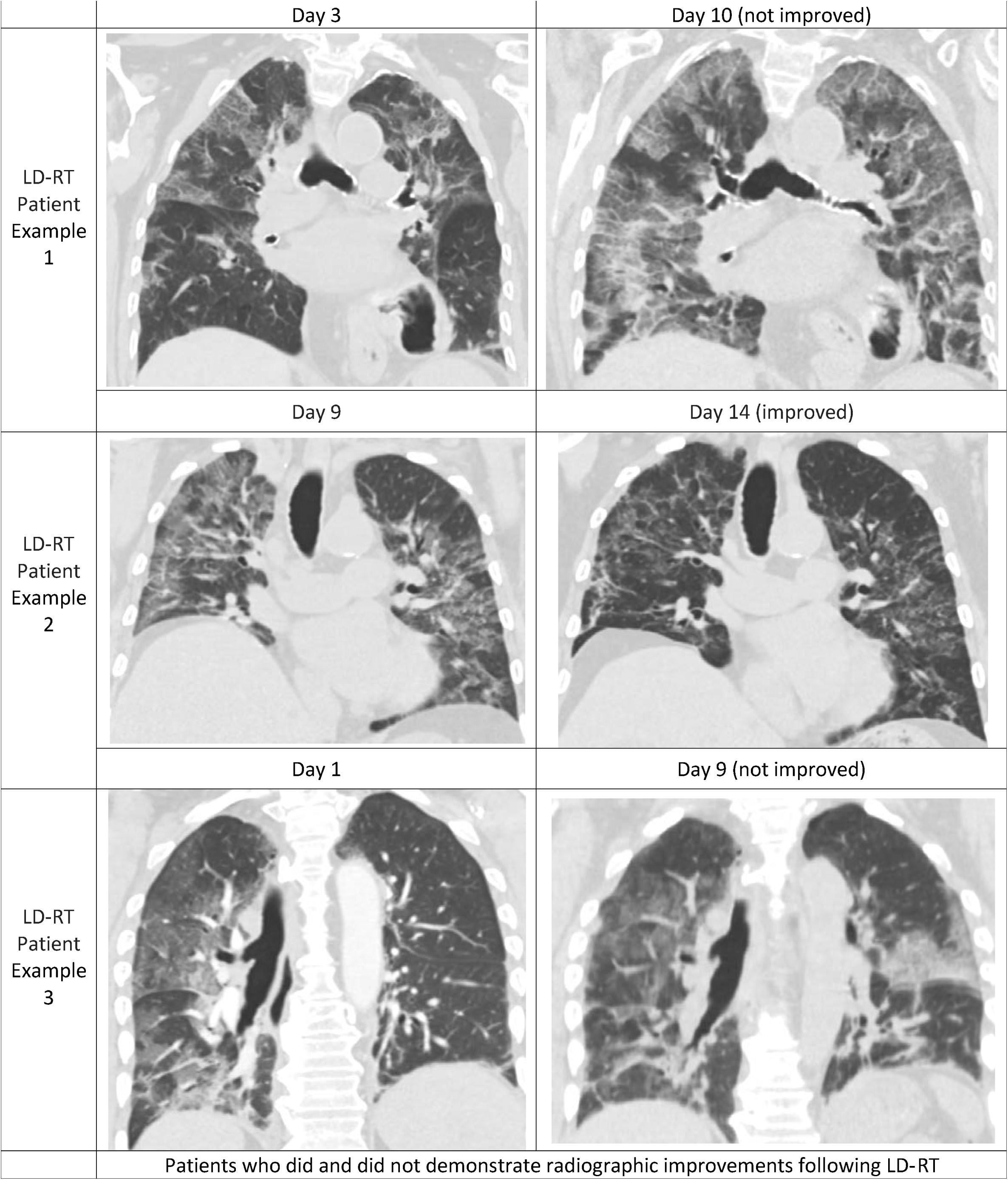
Radiographic Changes After Low-Dose Whole-Lung Radiotherapy (LD-RT)

**Figure S3.**
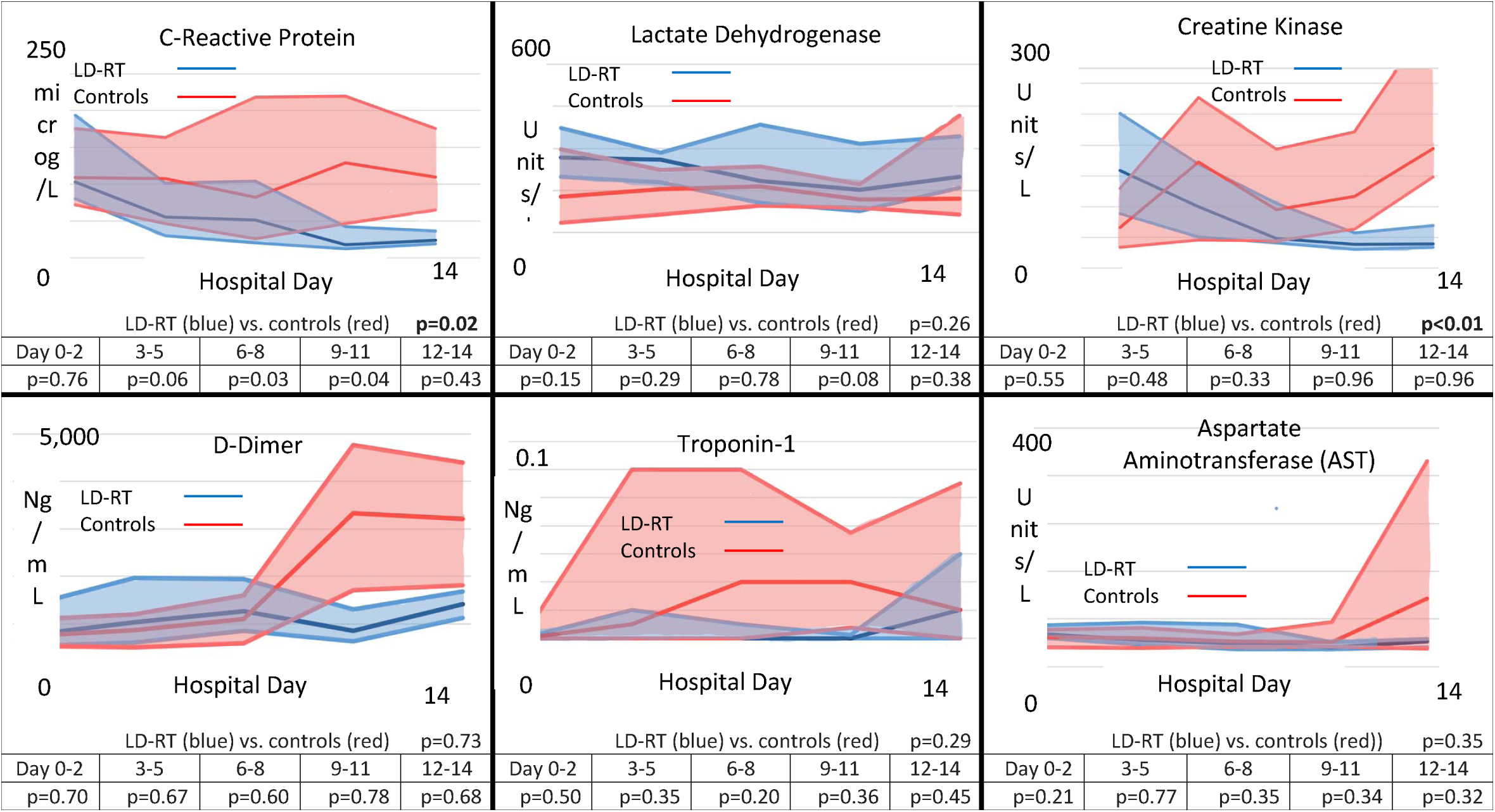
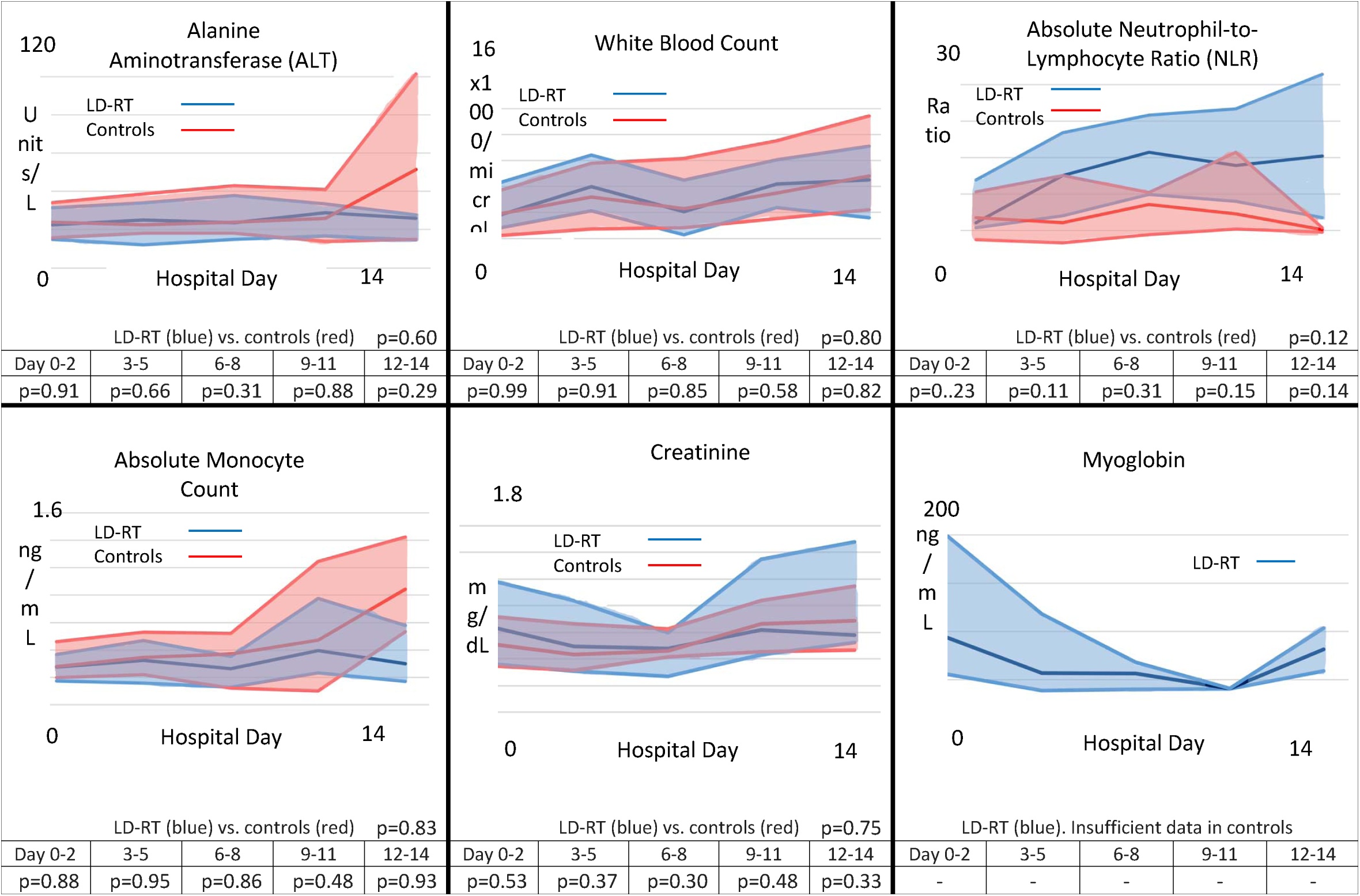
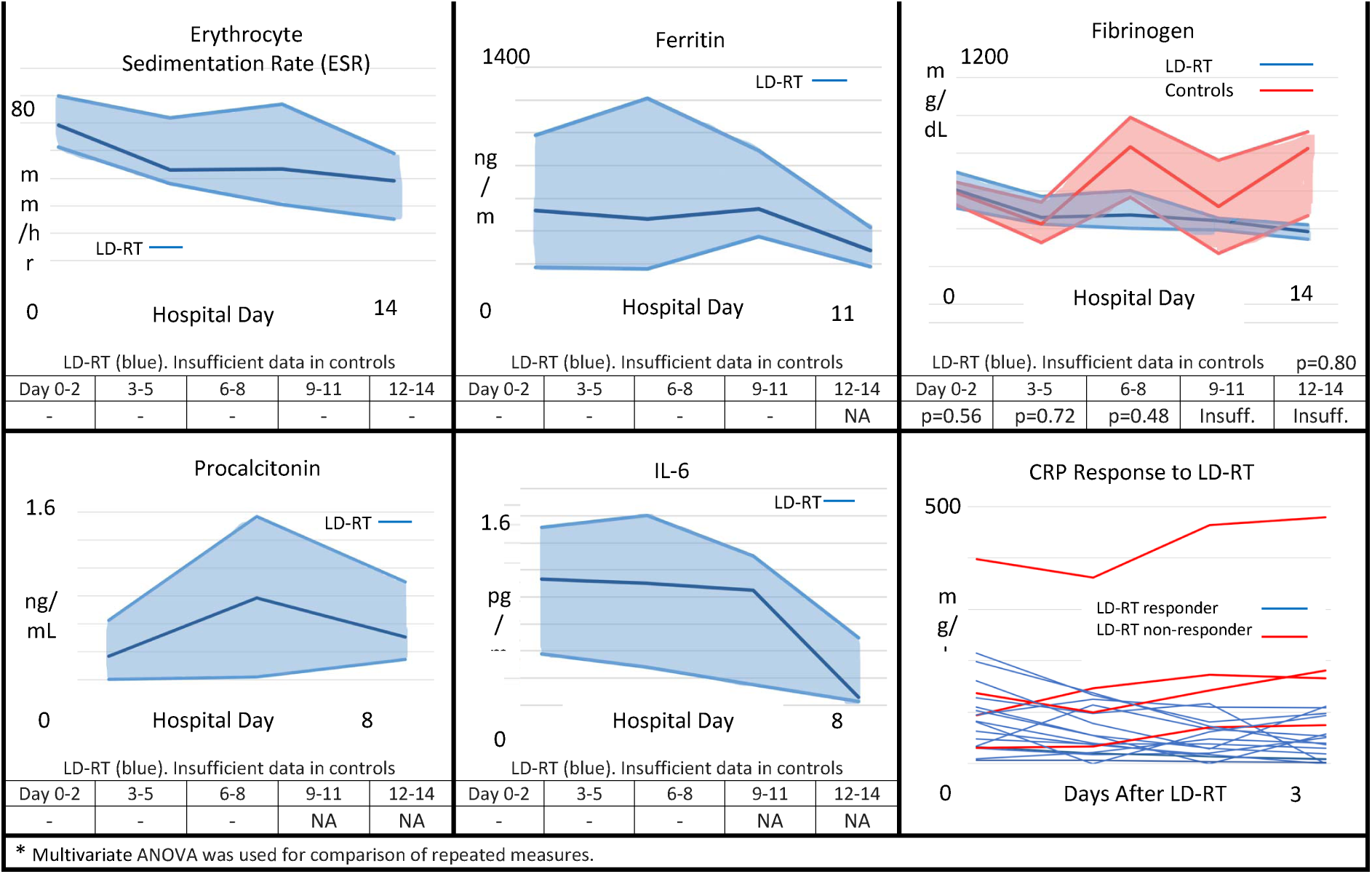
Serologic Median and Interquartile Ranges After Low-Dose Whole-Lung Radiation Therapy with Concurrent Dexamethasone and/or Remdesevir

## References

1. Mehta P, McAuley DF, Brown M, et al: COVID-19: consider cytokine storm syndromes and immunosuppression. Lancet 395:1033–1034, 2020

2. Huang C, Wang Y, Li X, et al: Clinical features of patients infected with 2019 novel coronavirus in Wuhan, China. Lancet 395:497–506, 2020

3. Bhatraju PK, Ghassemieh BJ, Nichols M, et al: Covid-19 in Critically Ill Patients in the Seattle Region - Case Series. N Engl J Med, 2020

4. Auld SC, Caridi-Scheible M, Blum JM, et al: ICU and Ventilator Mortality Among Critically Ill Adults With Coronavirus Disease 2019. Crit Care Med, 2020

5. Meziani L, Robert C, Classe M, et al: Low doses of radiation increase the immunosuppressive profile of lung macrophages during viral infection and pneumonia BIORXIV Online pre-print, 2020

6. Salomaa S, Cardis E, Bouffler SD, et al: Low dose radiation therapy for COVID-19 pneumonia: is there any supportive evidence? Int J Radiat Biol 96:1224–1227, 2020

7. Lara PC, Burgos J, Macias D: Low dose lung radiotherapy for COVID-19 pneumonia. The rationale for a cost-effective anti-inflammatory treatment. Clin Transl Radiat Oncol 23:27–29, 2020

8. Prasanna PG, Woloschak GE, DiCarlo AL, et al: Low-Dose Radiation Therapy (LDRT) for COVID-19: Benefits or Risks? Radiat Res 194:452–464, 2020

9. Hess CB, Nasti TH, Dhere V, et al: Immunomodulatory Low-Dose Whole-Lung Radiation for Patients with COVID-19-Related Pneumonia. Int J Radiat Oncol Biol Phys, 2020

10. Hess CB, Buchwald ZS, Stokes W, et al: Low-dose whole-lung radiation for COVID-19 pneumonia: Planned day 7 interim analysis of a registered clinical trial. Cancer 126:5109–5113, 2020

11. Ameri A, Rahnama N, Bozorgmehr R, et al: Low-Dose Whole-Lung Irradiation for COVID-19 Pneumonia: Short Course Results. Int J Radiat Oncol Biol Phys, 2020

12. Hanna CR, Robb KA, Blyth KG, et al: Clinician Attitudes to Using Low-Dose Radiation Therapy to Treat COVID-19 Lung Disease. Int J Radiat Oncol Biol Phys, 2020

13. Kirsch DG: Radiation Therapy as a Treatment for COVID-19? Int J Radiat Oncol Biol Phys 108:1140–1142, 2020

14. Khan MK, Hess CB: A Call to Action: “Low-Dose Radiation May Help Cure COVID-19…” [Taps Mic] “…Is This Thing On?”. JNCI Cancer Spectr 5:pkaa105, 2021

15. Charlson ME, Pompei P, Ales KL, et al: A new method of classifying prognostic comorbidity in longitudinal studies: development and validation. J Chronic Dis 40:373–83, 1987

16. Taylor E, Haven K, Reed P, et al: A chest radiograph scoring system in patients with severe acute respiratory infection: a validation study. BMC Med Imaging 15:61, 2015

17. Pan F, Ye T, Sun P, et al: Time Course of Lung Changes at Chest CT during Recovery from Coronavirus Disease 2019 (COVID-19). Radiology 295:715–721, 2020

18. Sharifpour M, Rangaraju S, Liu M, et al: C-Reactive protein as a prognostic indicator in hospitalized patients with COVID-19. PLoS One 15:e0242400, 2020

19. Lagunas-Rangel FA: Neutrophil-to-lymphocyte ratio and lymphocyte-to-C-reactive protein ratio in patients with severe coronavirus disease 2019 (COVID-19): A meta-analysis. J Med Virol 92:1733–1734, 2020

